# pyTCR: a comprehensive and scalable platform for TCR-Seq data analysis to facilitate reproducibility and rigor of immunogenomics research

**DOI:** 10.1101/2022.05.26.22275650

**Authors:** Kerui Peng, Jaden Moore, Jaqueline Brito, Guoyun Kao, Amanda M. Burkhardt, Houda Alachkar, Serghei Mangul

## Abstract

T cell receptor (TCR) studies have grown substantially with the advancement in the sequencing techniques of T cell receptor repertoire sequencing (TCR-Seq). The analysis of the TCR-Seq data requires computational skills to run the computational analysis of TCR repertoire tools. However biomedical researchers with limited computational backgrounds face numerous obstacles to properly and efficiently utilizing bioinformatics tools for analyzing TCR-Seq data. Here we report pyTCR, a computational notebook-based platform for comprehensive and scalable TCR-Seq data analysis. Computational notebooks, which combine code, calculations, and visualization, are able to provide users with a high level of flexibility and transparency for the analysis. Additionally, computational notebooks are demonstrated to be user-friendly and suitable for researchers with limited computational skills. Our platform has a rich set of functionalities including various TCR metrics, statistical analysis, and customizable visualizations. The application of pyTCR on large and diverse TCR-Seq datasets will enable the effective analysis of large-scale TCR-Seq data with flexibility, and eventually facilitate new discoveries.

## Introduction

T cell receptor (TCR) repertoire is a collection of all unique TCRs in an individual, which is formed through the process of V(D)J recombination after exposure to antigens and the activation of the adaptive immune response. With the growing understanding of TCR repertoire, researchers are able to leverage detailed TCR-Seq datasets to reveal the changes of TCR repertoires in a variety of human disease states such as cancer^1,2^, autoimmune diseases^3,4^, infectious diseases^5,6^, and neurodegenerative diseases^7,8^. Thus, these have helped the biomedical community to deepen the understanding of the roles of the adaptive immune system and adaptive immune responses. For example, studies have shown the usage and diversity of TCR repertoires could be utilized to help select the most suitable immunotherapy for cancer patients^9,10^. Thus, effective TCR profiling and analysis are informative to guide certain cancer treatments, which ultimately enables precision and personalized medicine.

With the rapid development of high-throughput sequencing techniques in the past decades, TCR-Seq has enabled researchers to effectively characterize TCR repertoires across various tissue types and diseases with high specificity and sensitivity by targeting TCR loci. Even with the available TCR profiling methods, TCR repertoire metrics such as diversity, gene usage, and motif enrichment cannot be easily interpreted directly from TCR-Seq data after initial TCR profiling. Post-analysis is required to calculate, visualize, and compare the sample level or population level TCR repertoire characteristics.

Existing bioinformatics tools for TCR repertoire post-analysis are available as R packages such as VDJTools^11^ and Immunarch^12^. These tools enable biomedical researchers to analyze TCR-Seq data, however, multiple barriers and limitations exist. First, as in any R package, users follow the instructions to enter commands in the command-line interface and the output will be presented in the summarized tables or figures. The analytical methods used for the particular step of the analysis are isolated, which can result in a limited understanding of the details of the analysis. This also increases the probability of human errors. Relying on the documentation of the tool is often not a reliable solution as it typically lacks details, has unclear and ambiguous wording, and can be outdated for future users. Second, the existing TCR-Seq analysis tools need to be installed and utilized with the command-line interface which can be a challenge for biomedical researchers who lack the required computational skill sets^13^. Third, the output files are generated as individual files. Biomedical researchers need to work between different files and tools to finish the post-analysis, which leads to high chances of creating manual mistakes. Last, none of the existing tools cover all aspects of the TCR-Seq analysis. For example, researchers need to use multiple packages or additional tools for statistical analysis, which adds an additional burden for biomedical researchers.

Here, we present pyTCR (python TCR analysis), an easy-to-use, interactive, and scalable platform with a wider range of functionalities compared to existing tools. pyTCR utilizes interactive computational notebooks to facilitate reproducibility and rigor of performed TCR-Seq analysis. The availability of well-documented code, visualization, and results of the analysis in a single notebook will facilitate transparency and reproducibility of performed analysis, make the users more aware of the details of the metrics and thresholds being used in the analysis, as well as minimize the possibility of manual mistakes and misinterpretation of the TCR-Seq data analysis results. Notably, pyTCR provides statistical analysis for the first time in a TCR-Seq analysis tool, which is not available in the existing tools. We have demonstrated the utility of pyTCR by applying it to the COVID19-BWNW dataset containing 46 TCR-Seq samples. Additionally, we have compared the scalability of our tool with the existing tools and have demonstrated substantial improvement in running time.

## Results

### pyTCR: a comprehensive and scalable platform for TCR-seq data analysis

pyTCR, an open-source, user-friendly platform that addresses the issues mentioned above, offers broader and more comprehensive TCR repertoire analysis with an increased number of types of analyses compared to the existing tools. Six types of analysis are contained in the pyTCR, which include basic analysis, clonality analysis, diversity analysis, overlap analysis, gene usage analysis, and motif analysis (Table 1). TCR repertoire metrics, visualization, and statistical analysis are included in all types of analyses (Fig.1, Table 1). Our platform for TCR-Seq data analysis uses interactive computational notebooks for post analysis and visualization of TCR-Seq data with a rich set of functionalities. Notably, we have used Google Colaboratory (Google Colab) to provide the cloud option, which is free to use and is a scalable option. No installation of the software to the local computers is required if the users choose to use the Google Colaboratory. The use of computational notebooks enables users to execute analysis and produce tabular output and customizable visualization so that users can use the desired features in their datasets to generate results.

**Table 1.** TCR repertoire analysis functions in pyTCR. We documented the name of the metrics (indicated in the column “Metric”) and the description of the corresponding metrics (indicated in the column “Description”).

| Metric | Description |
| --- | --- |
| Basic analysis |  |
| Read count | Number of reads in a sample |
| Clonotype count | Number of clonotypes in a sample, the diversity of VDJ combinations in a sample |
| Mean frequency | Mean of clonotype frequency in a sample<br>Sum of the clonotype frequency ÷ number of the clonotype |
| Geometric mean frequency | Geometric mean of clonotype frequency in a sample |
| Mean length of CDR3 nucleotide sequence | Mean length of CDR3 nucleotide sequence, weighted by clonotype frequency, in a sample<br>$\sum_{i=1}^n (len(nt) * pi)$<br>pi = frequency of clonotype i<br>n = number of unique clonotype in the sample<br>len(nt) = length of CDR3 nucleotide sequence |
| Convergence | Mean number of unique CDR3 nucleotide sequences that code for the same CDR3 amino acid sequence |
| Spectratype | Frequency of clonotypes based on CDR3 nucleotide lengths |
| Clonality analysis |  |
| The most or the least frequent clonotype | The most or the least frequent clonotype |
| 1-Pielou index | The evenness of the distribution of the clonotypes<br>$1 + \sum_{i=1}^n [(pi * \ln(pi)) / \ln(n)]$<br>pi = frequency of clonotype i<br>n = number of unique clonotype in the sample |
| Clonal proportion | The number of distinct clonotypes that accounts for greater than or equal to percentage (customizable) of the total of sequencing reads |
| Relative abundance (in all repertoire, top clonotypes, rare clonotypes) | The proportion of repertoire account for clonal groups with specific abundances in a sample |
| Diversity analysis |  |
| Shannon-Wiener index | $e^{\sum_{i=1}^n -(pi * \ln(pi))}$<br>pi = frequency of clonotype i<br>n = number of unique clonotype in the sample |
| Normalized Shannon-Wiener index | shannon-wiener index $\div \ln(n)$<br>n = number of unique clonotype in the sample |
| Inverse Simpson index | Simpson index $D = \sum_{i=1}^n pi^2$<br>1/D is the inverse simpson index |
| Gini Simpson index | 1-D is the gini simpson index |
| D50 index | The percentage of distinct clonotypes that accounts for greater than or equal to 50% of the total of sequencing reads |
| Chao1 estimate | Chao1 estimate = $S + \frac{f_1(f_1-1)}{2(f_2+1)}$<br>Variation of Chao1 estimate =<br>$f_2[0.5 * (\frac{f_1}{f_2})^2 + (\frac{f_1}{f_2})^3 + 0.25 * (\frac{f_1}{f_2})^4]$<br>f1 = number of clonotypes with read of 1<br>f2 = number of clonotypes with read of 2<br>S = number of clonotypes |
| Gini coefficient | The ratio of the area that lies between the line of equality and the Lorenz curve over the total area under the line of equality |
| Gene usage analysis |  |
| V,D,J gene weighted usage | Mean of gene frequency weighted by clonotype frequency |
| V,D,J gene unweighted usage | Mean of gene frequency by the number of the reads |
| Overlap analysis |  |
| Morisita-Horn index | $Ch = \frac{(2 * \sum_{i=1}^S x_i y_i)}{(\frac{\sum_{i=1}^S x_i^2}{X^2} + \frac{\sum_{i=1}^S y_i^2}{Y^2}) XY}$<br>x<sub>i</sub>: the number of times species i is represented in the total X from one sample<br>y<sub>i</sub>: the number of times species i is represented in the total Y from another sample |
|  | $D_x$ and $D_y$: the Simpson's index values for the x and y samples respectively<br>S: the number of unique species |
| Jaccard index | Jaccard Similarity = (number of observations in both sets) / (number in either set) |
| Overlap coefficient | The size of the intersection divided by the smaller of the size of the two sets |
| Tversky index | $\frac{BothAB}{\alpha * onlyA + \beta * onlyB + bothAB}$<br>$\alpha = \beta = 0.5$ (Sørensen–Dice coefficient) |
| Cosine similarity | $\frac{\sum_{i=1}^n A_i B_i}{\sqrt{\sum_{i=1}^n A_i^2} \sqrt{\sum_{i=1}^n B_i^2}}$ |
| Pearson correlation of clonotype frequencies | $R_{ij} = \frac{\sum_{k=1}^N (\phi_{ik} - \bar{\phi}_i)(\phi_{jk} - \bar{\phi}_j)}{\sqrt{\sum_{k=1}^N (\phi_{ik} - \bar{\phi}_i)^2} \sqrt{\sum_{k=1}^N (\phi_{jk} - \bar{\phi}_j)^2}}$<br>N: the index of overlapping clonotypes<br>$\phi_{ik}$: the frequency of clonotype k in sample i |
| Relative overlap diversity | $D_{ij} = \frac{d_{ij}}{d_i * d_j}$<br>$d_{ij}$: the number of clonotypes present in both samples<br>$d_i$: the diversity of sample i |
| Geometric mean of relative overlap frequencies | $F_{ij} = \sqrt{f_{ij} * f_{ji}}$<br>$f_{ij} = \sum_{k=1}^N \phi_{ik}$: the total frequency of clonotypes that overlap between samples i and j in sample i |
| Clonotype-wise sum of geometric mean frequencies | $F2ij = \sum_{k=1}^N \sqrt{\phi_{ik}\phi_{jk}}$<br>$\phi_{ik}$: the frequency of clonotype k in sample i |
| Jensen-Shannon divergence | $JS(P, Q) = \frac{1}{2}(KL(P, \frac{P+Q}{2}) + KL(Q, \frac{P+Q}{2}))$ $KL(P, Q) = \sum_i p_i \log_2 \frac{p_i}{q_i}$ |
| Motif analysis |  |
| Amino acid spectratype | Frequency of clonotypes based on CDR3 amino acid lengths |
| Amino acid motif analysis | Counts of the amino acid motifs |
| Nucleotide sequence motif analysis | Counts of the nucleotide sequence motifs |

**Figure 1.**
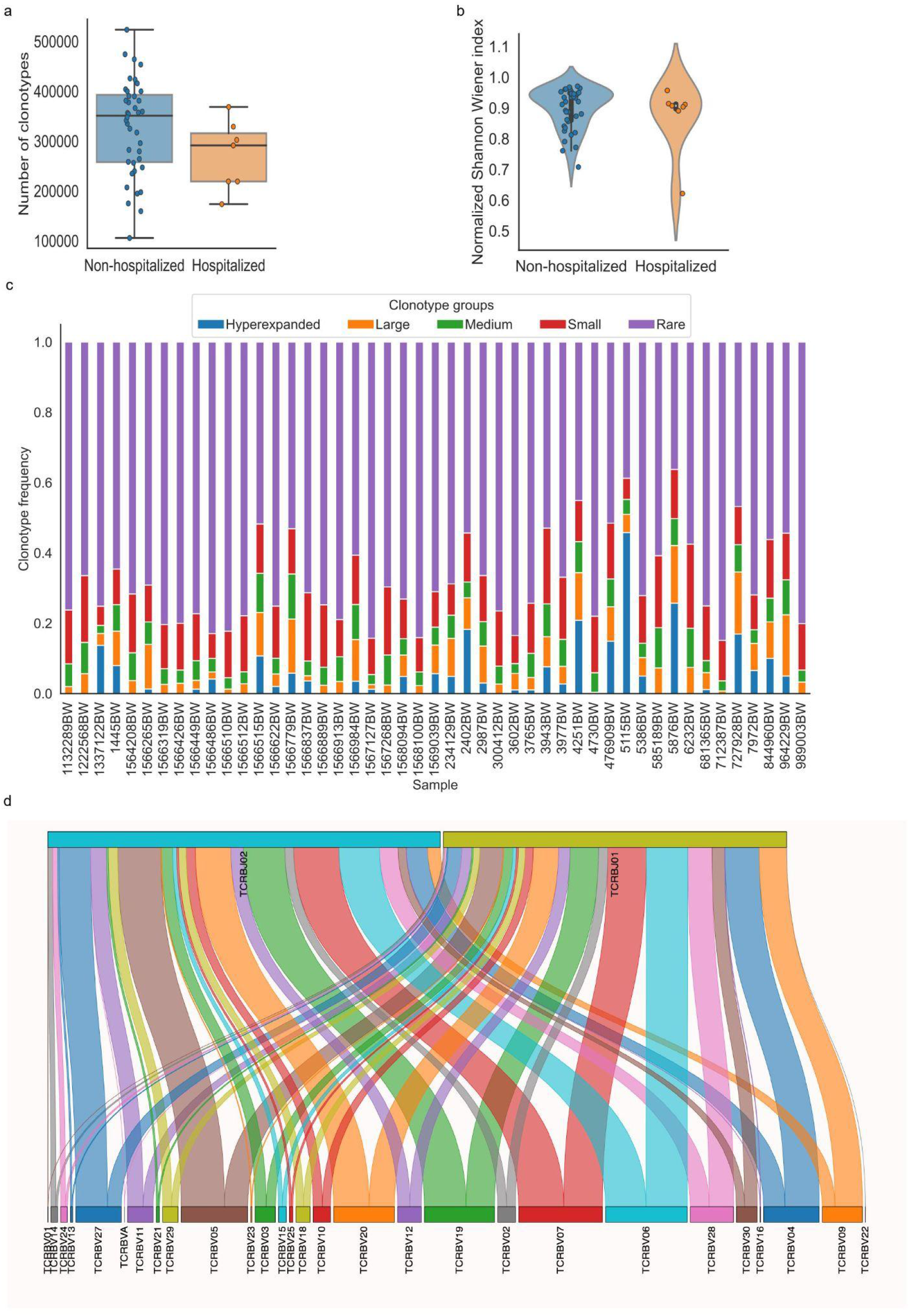
Visualization of TCR repertoire metrics generated using pyTCR. a. The clonotype counts of each sample grouped by hospitalization status were presented as a box plot and strip plot. b. The normalized Shannon-Wiener index of each sample grouped by hospitalization status was presented as a violin plot. c. The distribution of clonotype groups in each sample was presented as a stacked bar plot. The clonotypes were categorized into five groups based on the clonotype frequencies. Hyperexpanded clonotypes were the clones with frequencies between 0.01 to 1, large clonotypes were the clones with frequencies between 0.001 to 0.01, medium clonotypes were the clones with frequencies between 0.0001 to 0.001, small clonotypes were the clones with frequencies between 0.00001 to 0.0001, rare clonotypes were the clones with frequencies between 0 to 0.00001. d. The V-J combinations with V and J gene frequencies in sample 1445BW were presented as a Sankey plot.

The users can clone the GitHub repository to their local computer and utilize the pyTCR notebooks locally via Jupyter Notebook^14^. Users who choose to use Google Colab (cloud option of pyTCR) can upload the data files from local computers, web-based drives, or GitHub repositories. The results can be downloaded and stored locally or into web-based drives by the code provided. pyTCR accepts inputs from pre-processing software including MiTCR, MiXCR, and ImmunoSEQ. The minimal clonotype information should include the counts of reads, clonotype frequency, CDR3 nucleotide sequence, CDR3 amino acid sequence, and the inferred V, D, and J genes in the input data files. pyTCR provides conversion to the corresponding format which consists of the columns of counts of reads (#count), frequency (freq), CDR3 sequence (cdr3nt), CDR3 amino acids (cdr3aa), V gene (v), D gene (d), J gene (j), features (if provided), sample name. These should be already filtered for the non-coding CDR3 by the upstream tools. If the metadata is not available, a notebook for combining individual files to a metadata file should be executed prior to any TCR-Seq analysis to reduce the burden of analyzing individual data files separately. In order to achieve this, all the sample files should be stored or uploaded in one folder prior to generating the metadata file. The notebook that combines individual sample data files to a metatable with all the files is provided.

### pyTCR is able to perform basic analysis to characterize the TCR repertoire

The focus of the basic analysis is to group and provide the most fundamental TCR repertoire metrics in one place. The basic analysis performed by pyTCR estimates provides the number of reads, clonotype counts, mean clonotype frequency, the geometric mean of clonotype frequency, mean length of CD3 nucleotide sequence, convergence, spectratype as TCR repertoire metrics. The visualization is available for all the metrics (except for spectratype) in the basic analysis at the individual sample level and group level. The available plot types are violin plot, strip plot, swarm plot, box plot, boxen plot, point plot, and bar plot (Fig.1a, Supplementary Fig.1). We were able to detect that the mean reads count in the hospitalization group was lower than that in the non-hospitalization group (480844.1 and 554580.3, respectively; t-test: p = 0.229), and the mean clonotype count in the hospitalization group was lower than in the non-hospitalization group (271777.6 and 328980.1, respectively; t-test: p = 0.136) in the COVID19-BWNW dataset.

### pyTCR is able to perform clonality analysis to assess the evenness of distribution of TCR clonotypes

The clonality analysis offers the measurements of clonality, which has been used to assess the evenness of distribution of the clonotypes based on the relative abundance of clonotypes in the sample. The metrics include the list of the most or the least frequent clonotypes, 1-Pielou index for evenness measure (0 means no evenness, 1 means complete evenness), clonal proportion, and the distribution of clonotype groups based on relative abundance. Specifically, clonal proportion presents the number of clonotypes that consist of a certain percentage of the clonotypes in the repertoire. In the COVID19-BWNW dataset, the number of clonotypes that counts for 10% of the clonotypes in the repertoire was smaller in the hospitalization group than in the non-hospitalization group (49.5 and 459, respectively; Wilcoxon rank-sum test, p = 0.596), the corresponding plots were presented in various types (Supplementary Fig.2). Additionally, the distribution of clonotype groups based on clonotype frequency or count in each sample can be presented in bar plots across all the clonotypes, the top clonotypes, and the rare clonotypes (Supplementary Fig.3). We presented the distribution of five clonotype groups (hyperexpanded, large, medium, small, and rare) across all clonotypes in Fig.1c, this categorization is similarly done in other existing tools. The users have full control of the thresholds of the clonotype groups in our tool.

### pyTCR is able to perform gene usage analysis to detect over and underrepresented TCR genes across the samples

Gene usage analysis provides the weighted and unweighted V/D/J gene usage calculations. For gene usage analysis, V gene usage, D gene usage, and J gene usage, both weighted (which is based on clonotype frequency) and unweighted (which is based on clonotype count) are provided as TCR repertoire metrics. Heatmap and hierarchically clustered heatmap are the available visualizations (Supplementary Fig.4a-b). Sankey plot is also available to visualize the V-J combinations (Fig.1d, Supplementary Fig.4c), this is not provided by other existing tools. We observed higher V gene weighted usage of TRBV05-05*01 (0.0084 and 0.0066, respectively) and TRBV13-01*01 (0.0069 and 0.0042, respectively) in the non-hospitalization group. In comparison, we observed higher V gene weighted usage of TRBV20 (0.0638 and 0.0588, respectively) in the hospitalization group in the COVID19-BWNW dataset. We also observed higher V gene unweighted usage of TRBV18-01*01 (0.035 and 0.031) and TRBV30-01*01 (0.025 and 0.019) in the hospitalization group. After the Bonferroni correction to account for the multiple comparisons, according to the adjusted p values, the differences mentioned above were not statistically significant.

### pyTCR is able to assess the diversity of TCR repertoires

Diversity analysis offered by pyTCR includes all the widely adopted indices to characterize the diversity of TCR repertoire, which contains Shannon-Wiener index, normalized Shannon-Wiener index, inverse Simpson index, Gini Simpson index, D50 index, Chao1 estimate, Gini coefficient (Table 1). High Shannon-Wiener index, low normalized Shannon-Wiener index, high inverse Simpson index, high Gini Simpson index, high Chao1 estimate, and high Gini coefficient represent high clonal diversity. Additionally, the D50 index represents the percentage of unique clonotypes that account for greater than 50% of the total number of sequences. The visualization is available for all the diversity metrics at the sample or group level as violin plot, strip plot, swarm plot, box plot, boxen plot, point plot, and bar plot (Fig.1b, Supplementary Fig.5). In the COVID19-BWNW dataset, the median Shannon-Wiener index, the median inverse Simpson index, and the median Gini Simpson index were all lower in the hospitalization group than in the non-hospitalization group. Even though none of the diversity indices was statistically significant, most of the diversity indices showed the trend that patients in the non-hospitalization group have more diverse TCR clonotypes than patients in the hospitalization group. This finding was consistent with the results observed in the previously published studies, that severe COVID-19 patients had reduced TCR diversity than moderate COVID-19 patients^15,16^.

### pyTCR is able to effectively compare clonotypes and motifs across samples

The overlap analysis offers a comprehensive list of overlap metrics for comparing the clonotype frequencies between two samples. These metrics include the Jaccard index, overlap coefficient, Morisita-Horn index, Tversky index, Cosine similarity, Pearson correlation of clonotype frequencies, relative overlap diversity, the geometric mean of relative overlap frequencies, the clonotype-wise sum of geometric mean frequencies, and Jensen-Shannon divergence. For overlap analysis, the visualization is shown in the heatmaps (Supplementary Fig.6). Currently, existing tools only accept one sample per file for overlap comparisons which can be difficult to manage if the data already contains multiple samples per file. pyTCR allows for an unlimited amount of samples per file which enables more flexibility and less file management.

Motif analysis provides enriched nucleotide and amino acid motif discovery with customized length. For motif analysis, the amino acid motif counts and nucleotide motif counts in each sample are provided. The users are able to customize the length of the motif and visualize the distribution of the motifs in each sample or each group. In the COVID19-BWNW dataset, we observed amino acid motifs NTEAFF, YNEQFF, CASSLG, TDTQYF, NQPQHF, TGELFF, SYEQYF were the most abundant ones in both the hospitalization and non-hospitalization groups (Supplementary Fig.7a). We also observed nucleotide motifs such as TCTGTG, CTGTGC, TGTGCC, GTGCCA, TGCCAG, GCCAGC, CCAGCA, CAGCAG were the most abundant ones in both hospitalization and non-hospitalization groups (Supplementary Fig.7b).

### pyTCR offers several advantages compared to the existing tools

pyTCR provides more comprehensive functionalities compared with VDJtools and Immunarch (Supplementary Table 1). Notably, pyTCR includes several innovations that have not been implemented in the VDJtools or Immunarch before. First, statistical analyses for TCR-Seq datasets are embedded in pyTCR computational notebooks. No additional software or platform is needed for statistical analysis. Second, pyTCR has the most comprehensive list of overlap indices, which enable the thorough comparison between genotypes across samples. Third, pyTCR offers enriched motif detection for both amino acid sequences as well as nucleotide sequences. Furthermore, we have compared the results produced by pyTCR with VDJtools on the types of analysis that are provided by VDJtools on the COVID19-BWNW dataset. The results are consistent across our tool and VDJtools.

We also evaluated the scalability of pyTCR by varying the number of samples in the TCR-Seq data input files. After subsampling the COVID19-BWNW dataset into data files containing 2 to 46 samples, we recorded the central processing unit (CPU) time and memory usage required by pyTCR, VDJtools, and Immunarch when running overlap analysis. We observed that pyTCR required a significantly less amount of CPU time across all the subsamples compared to VDJtools and Immunarch (Fig.2a). For example, on average, pyTCR used 5 minutes and 25 seconds to process the overlap analysis for 46 samples, while Immunarch used 1 hour 49 minutes and 46 seconds to process the same number of samples. In terms of memory usage, pyTCR had reduced memory usage for most of the subsamples (Fig.2b). According to our benchmark results, we observe that pyTCR has up to 22 times faster performance than existing TCR-seq analysis tools, especially for datasets with larger numbers of samples.

**Figure 2.**
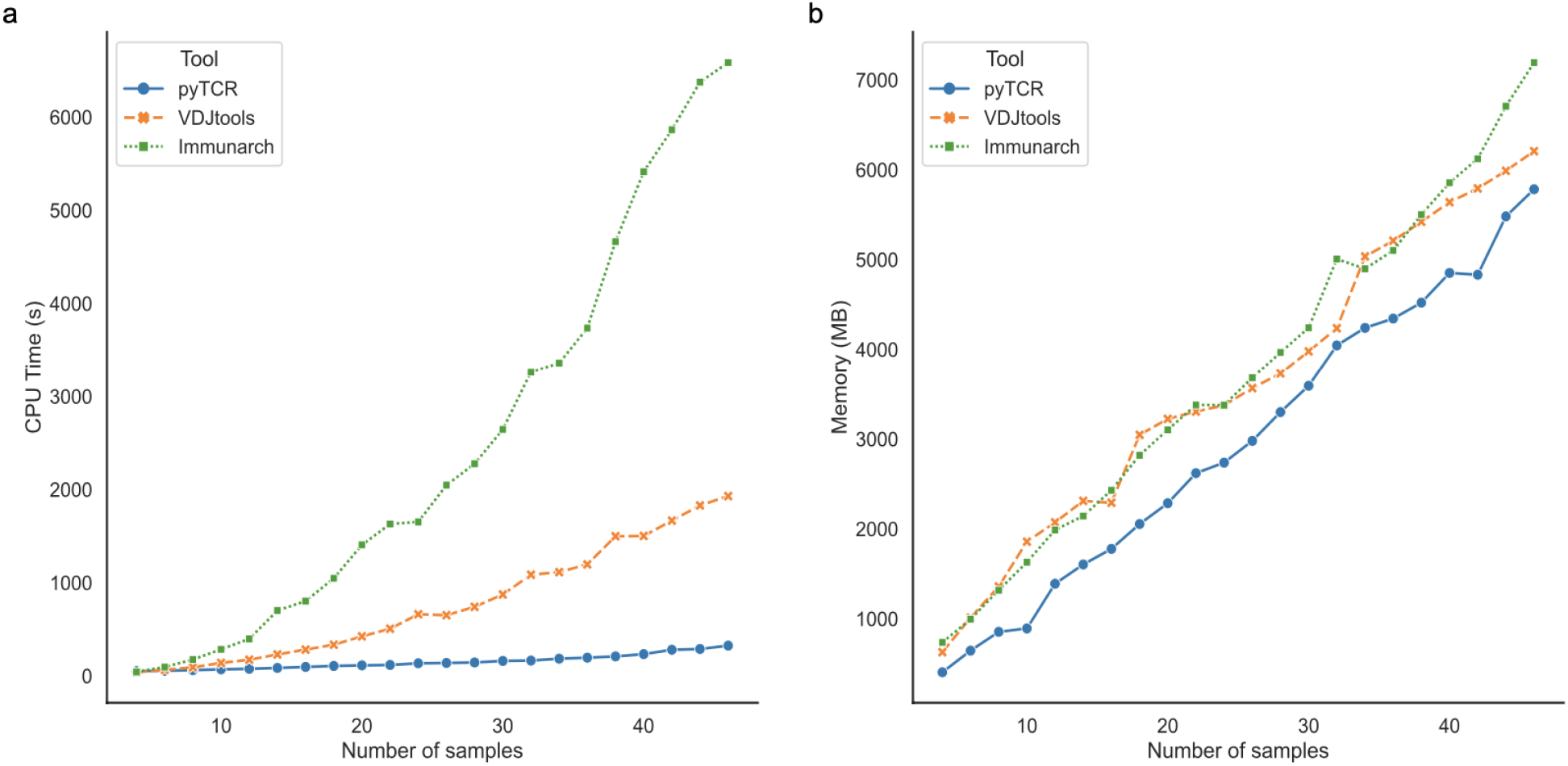
Central Processing Unit (CPU) time (a) and Memory usage (b) for subsamples of the COVID19-BWNW dataset for overlap analysis. Each point on the line plot represented the average of 10 runs for different input sizes.

## Discussion

We have presented pyTCR, a comprehensive and scalable computational notebook-based platform for TCR-Seq analysis and visualization with a rich set of functionalities. For the cloud-based version, we use Google Colaboratory (Google Colab). Google Colab, as a user-friendly, free with no installation needed prior to use service for Google account holders, is suitable for biomedical researchers with a limited computational background. Using interactive computational notebooks promotes high transparency for biomedical researchers because the steps of analyzing and visualizing are recorded and saved, which are easy to be shared with the scientific community.

pyTCR offers several advantages compared to the existing tools. First, pyTCR includes more comprehensive measurements than existing tools to analyze TCR-Seq data. The enriched measurements can provide users with more options to effectively characterize TCR repertoires and compare across various phenotypes. Furthermore, pyTCR provides code and analysis jointly together. Users can understand the definition of measurements and interpret results easily with pyTCR, as the explanation of the code and the math equations are available in the notebooks. Additionally, pyTCR allows users to adjust parameters easily and directly in the notebooks. Unlike other traditional bioinformatics tools, changing parameters that generate separate files which leads to high error rates by analyzing across different files, pyTCR provides all the analysis to be performed in the cloud where the files are automatically saved with the updated parameters and no generation of different files is needed. Last, our tool is more scalable as it requires less computational time for analysis.

In conclusion, our tool offers broader and more powerful functions in TCR repertoire research. We expect the computational notebook-based platform to be adopted by the broad biomedical community as it carries benefits that are superior or comparable to R packages.

## Method

### TCR-Seq data

We used the COVID19-BWNW dataset from the Adaptive ImmuneRace study to demonstrate the functionality of pyTCR. COVID19-BWNW dataset contains 46 convalescent COVID-19 patient samples collected at Bloodworks Northwest. Demographic and clinical features including age, gender, smoking status, ICU admit status, birth year, blood type, CMV at donation, days from the last symptom to sample date, ethnicity, race, height, weight, and hospitalization status are reported. The extracted genomic DNA was sequenced based on Multiplex PCR and only for the TCR beta chain by using the MiSeq platform.

### TCR-Seq data preprocessing

We downloaded 46 TCR-Seq data samples and the file containing demographic and clinical features in the tab-separated values (tsv) format. All the demographic and clinical features were listed in the sample_tags column in the file. The features were split into one in each column for further analysis.

### Statistical analysis

The statistical analysis is available for comparing numerical values across two groups. We first examine whether the datasets are normally distributed. If the dataset is normally distributed, we use the student’s t-test to evaluate the statistical significance. Otherwise, we use Wilcoxon rank-sum test to evaluate the statistical significance. Bonferroni correction is also included to count for the multiple comparisons across different genes.

### Jupyter notebooks

Jupyter notebooks (https://jupyter.org) is a web-based interactive computing platform that contains code, markdown text, and visualizations. These features enable users to conduct reproducible and transparent data analysis. We develop pyTCR based on Jupyter notebooks. In each Jupyter cell, we include code for either calculation for analysis or visualization. Users can easily change the parameters in the code to generate the results of their interests. Markdown text is used for instructions and explanations.

### pyTCR data structure and software implementation

The individual input data file can be text files, tab-separated values (tsv) files, or comma-separated values (csv) files. pyTCR uses a small suite of robust open-source python libraries to facilitate complex data analysis and visualizations. Python libraries including Pandas, Numpy, Matplotlib, Seaborn, and Scipy are used to provide rich functionality with limited amounts of code. Pandas is used to convert tabularly formatted TCR-Seq data into python data frames for notebooks to utilize. Numpy is used to perform complex mathematical operations across python data frames. Matplotlib and Seaborn are then used in tandem to generate rich data visualizations from the resultant data.

One critical component of pyTCR functionality is the overlap analysis between two samples. Such operations are unavoidably expensive in terms of computing power. Yet, pyTCR employs multiple different technical optimizations by default to provide the most optimal performance for researchers. In the case of any piecewise comparison between two samples, we first index and group the data into a key-value pair hash table for instantaneous look-up time. We then uniquely merge samples for comparison and index them using a hash table in the same manner. By utilizing this method, we are able to negate a large amount of computing time that would otherwise be associated with searching for the correct samples in the data set. This method allows us to instantly retrieve the required samples for future look-ups which shifts most of the computing time from slow searches, back onto the piecewise comparisons.

### Comparison with other methods

We used the COVID19-BWNW dataset to compare pyTCR to VDJtools and Immunarch for benchmarking purposes. We subsampled the COVID19-BWNW dataset to files containing 2, 4, 6, 8, 10, 12, 14, 16, 18, 20, 22, 24, 26, 28, 30, 32, 34, 36, 38, 40, 42, 44, 46 TCR-Seq samples. We then ran the overlap analysis of each tool ten times and computed the average CPU time and RAM usage for each. For the comparison, we utilized a high performance computing cluster (HPCC) to acquire the most accurate benchmarking results. However, in the comparison of 2 TCR-Seq samples for VDJtools and Immunarch, the HPCC was unable to record the results due to the short nature of the task. That is, the benchmark ended too quickly for the HPCC to accurately record the results. Thus the results for 2 TCR-Seq samples were not taken with the average of ten runs. Instead, we recorded the results once by introducing an artificial stall in the benchmark such that the HPCC had time to record, and then we subtracted the artificial stall time from the final CPU time. The RAM usage remains unchanged with this workaround.

## Data Availability

All data produced are available online at https://github.com/Mangul-Lab-USC/pyTCR.

## Data availability

COVID19-BWNW dataset was part of the ImmuneRACE Study, which was downloaded from ImmuneCODE™ database (https://immunerace.adaptivebiotech.com/data). Each individual data file is available at https://github.com/Mangul-Lab-USC/pyTCR. All processed and summarized data required to produce the figures and analysis performed in this paper are available at https://github.com/Mangul-Lab-USC/pyTCR, including the data used to produce all figures.

## Code availability

The code that was used to produce the figures and analysis performed in this paper are available at https://github.com/Mangul-Lab-USC/pyTCR.

## Acknowledgments

We would like to thank Yesha Patel for the helpful suggestions for statistical analysis. KP and SM are supported by the National Science Foundation grants 2041984 and 2135954. The authors acknowledge the Center for Advanced Research Computing (CARC) at the University of Southern California for providing computing resources that have contributed to the research results reported within this publication. URL: https://carc.usc.edu.

## Supplementary Tables

**Supplementary Table 1.**
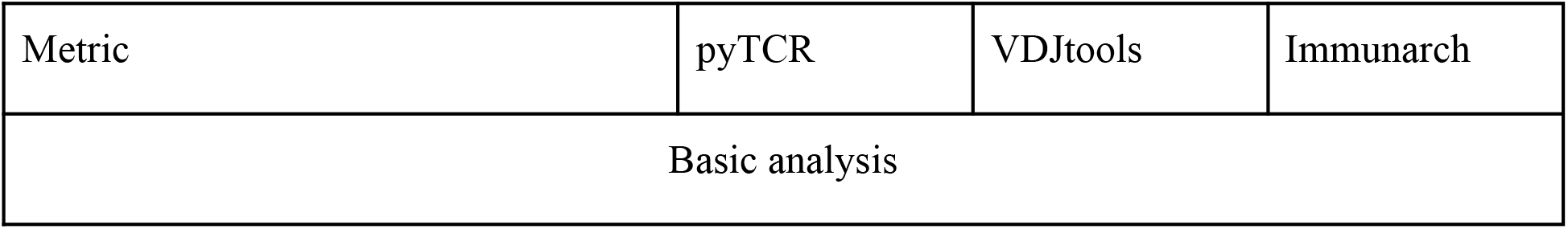
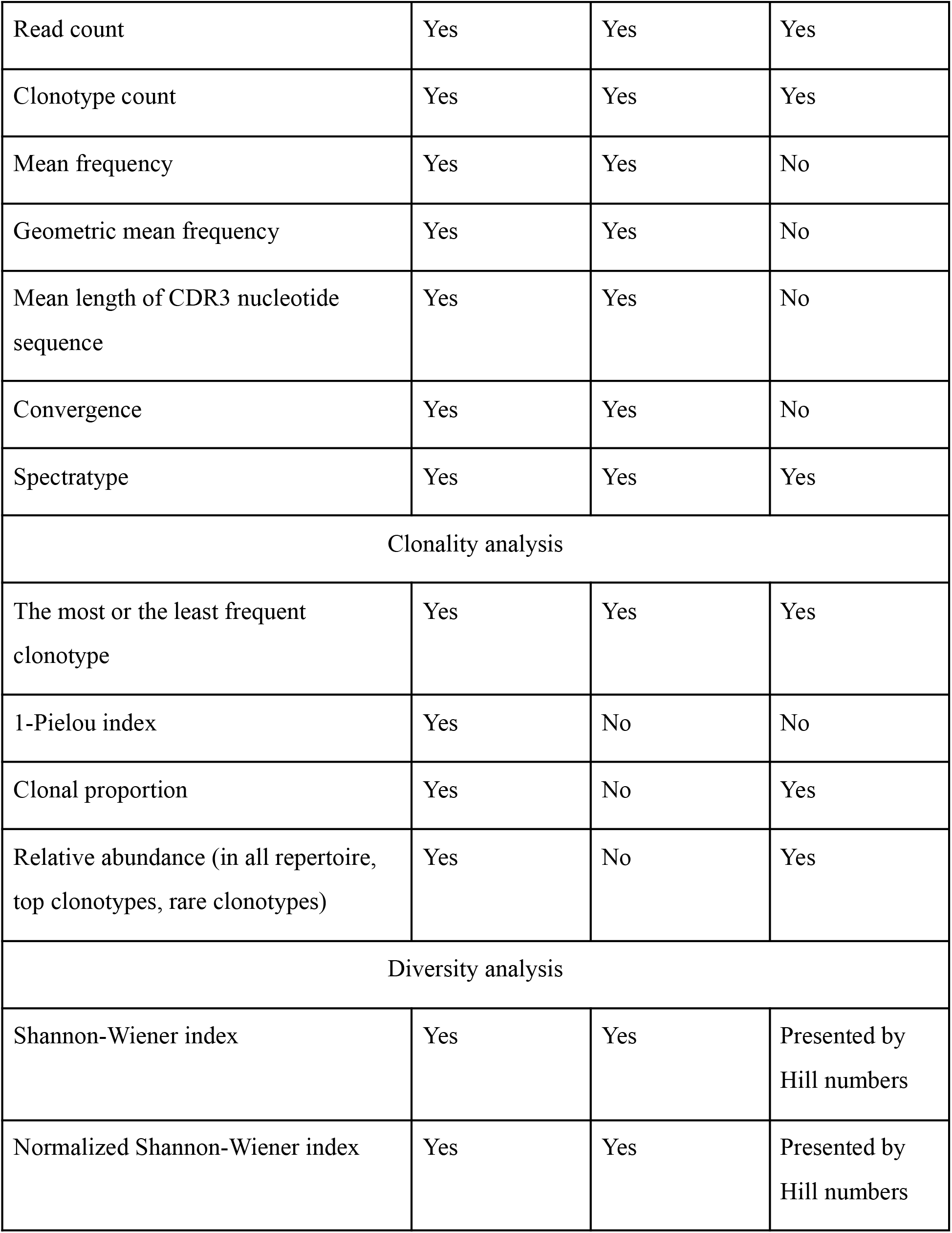
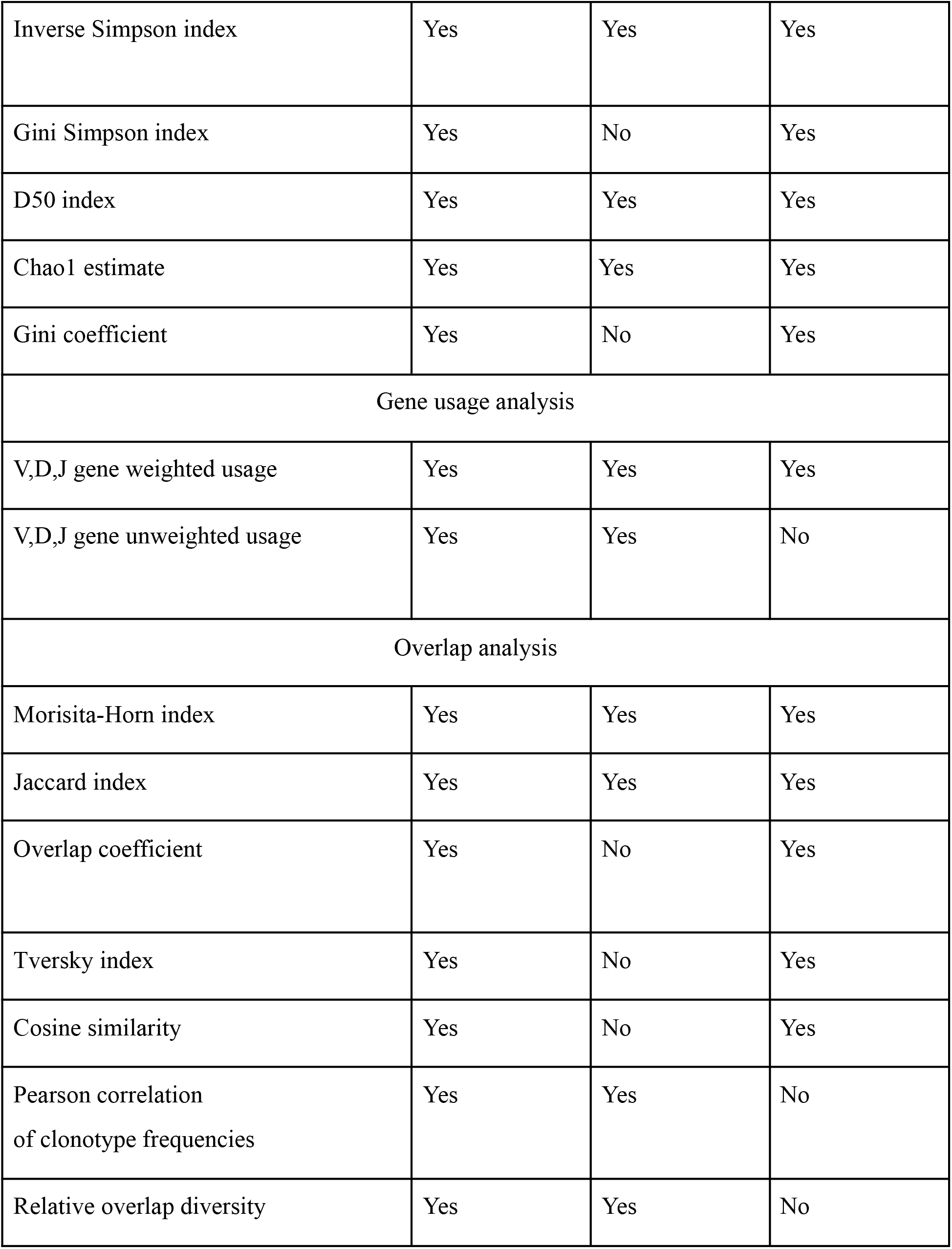
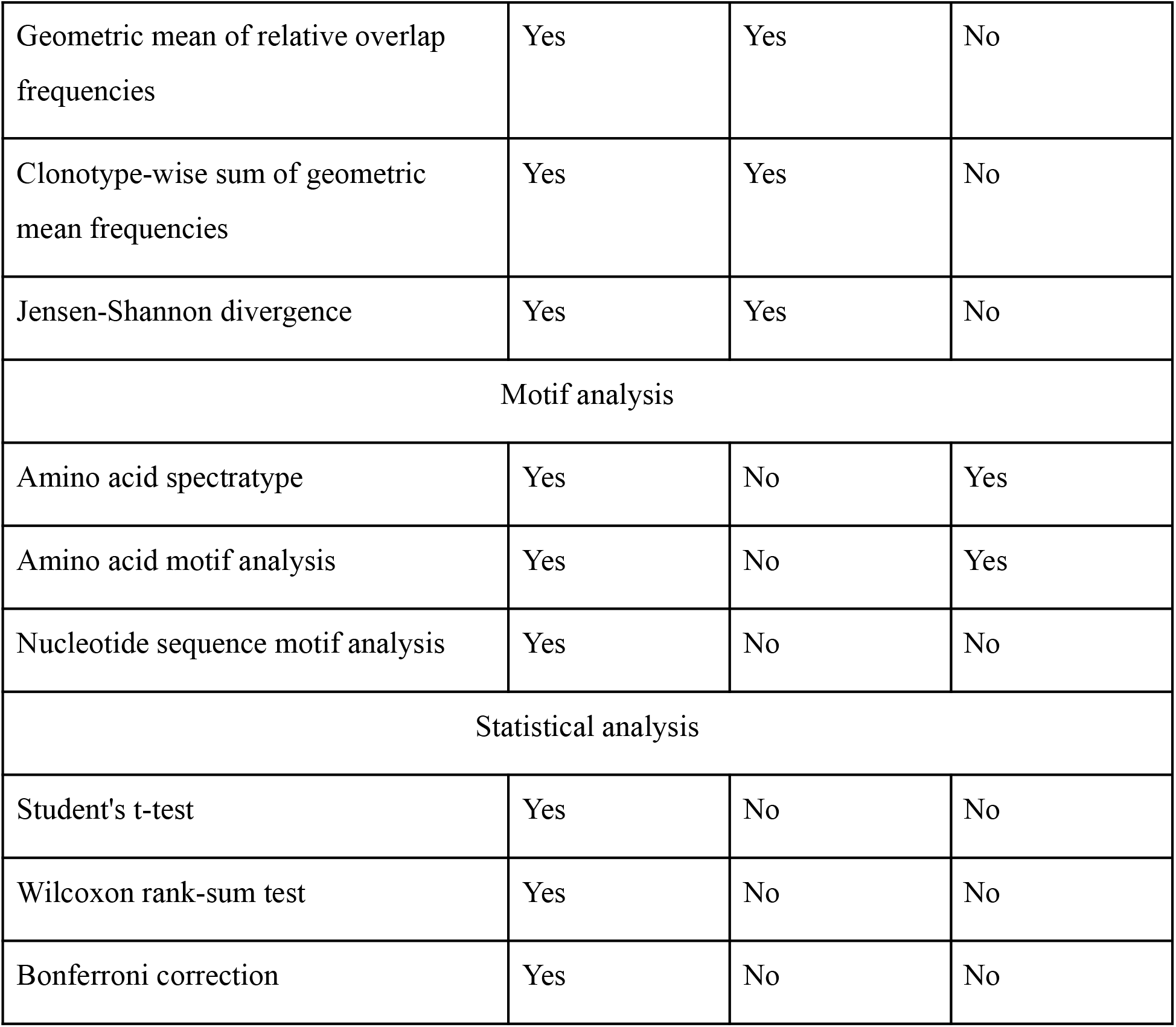
The comparison of TCR repertoire analysis tools.

| Metric | pyTCR | VDJtools | Immunarch |
| --- | --- | --- | --- |
| Basic analysis |  |  |  |
| Read count | Yes | Yes | Yes |
| Clonotype count | Yes | Yes | Yes |
| Mean frequency | Yes | Yes | No |
| Geometric mean frequency | Yes | Yes | No |
| Mean length of CDR3 nucleotide sequence | Yes | Yes | No |
| Convergence | Yes | Yes | No |
| Spectratype | Yes | Yes | Yes |
| Clonality analysis |  |  |  |
| The most or the least frequent clonotype | Yes | Yes | Yes |
| 1-Pielou index | Yes | No | No |
| Clonal proportion | Yes | No | Yes |
| Relative abundance (in all repertoire, top clonotypes, rare clonotypes) | Yes | No | Yes |
| Diversity analysis |  |  |  |
| Shannon-Wiener index | Yes | Yes | Presented by Hill numbers |
| Normalized Shannon-Wiener index | Yes | Yes | Presented by Hill numbers |
| Inverse Simpson index | Yes | Yes | Yes |
| Gini Simpson index | Yes | No | Yes |
| D50 index | Yes | Yes | Yes |
| Chao1 estimate | Yes | Yes | Yes |
| Gini coefficient | Yes | No | Yes |
| Gene usage analysis |  |  |  |
| V,D,J gene weighted usage | Yes | Yes | Yes |
| V,D,J gene unweighted usage | Yes | Yes | No |
| Overlap analysis |  |  |  |
| Morisita-Horn index | Yes | Yes | Yes |
| Jaccard index | Yes | Yes | Yes |
| Overlap coefficient | Yes | No | Yes |
| Tversky index | Yes | No | Yes |
| Cosine similarity | Yes | No | Yes |
| Pearson correlation<br>of clonotype frequencies | Yes | Yes | No |
| Relative overlap diversity | Yes | Yes | No |
| Geometric mean of relative overlap frequencies | Yes | Yes | No |
| Clonotype-wise sum of geometric mean frequencies | Yes | Yes | No |
| Jensen-Shannon divergence | Yes | Yes | No |
| Motif analysis |  |  |  |
| Amino acid spectratype | Yes | No | Yes |
| Amino acid motif analysis | Yes | No | Yes |
| Nucleotide sequence motif analysis | Yes | No | No |
| Statistical analysis |  |  |  |
| Student's t-test | Yes | No | No |
| Wilcoxon rank-sum test | Yes | No | No |
| Bonferroni correction | Yes | No | No |

## Supplementary Figures

**Supplementary Fig.1.**
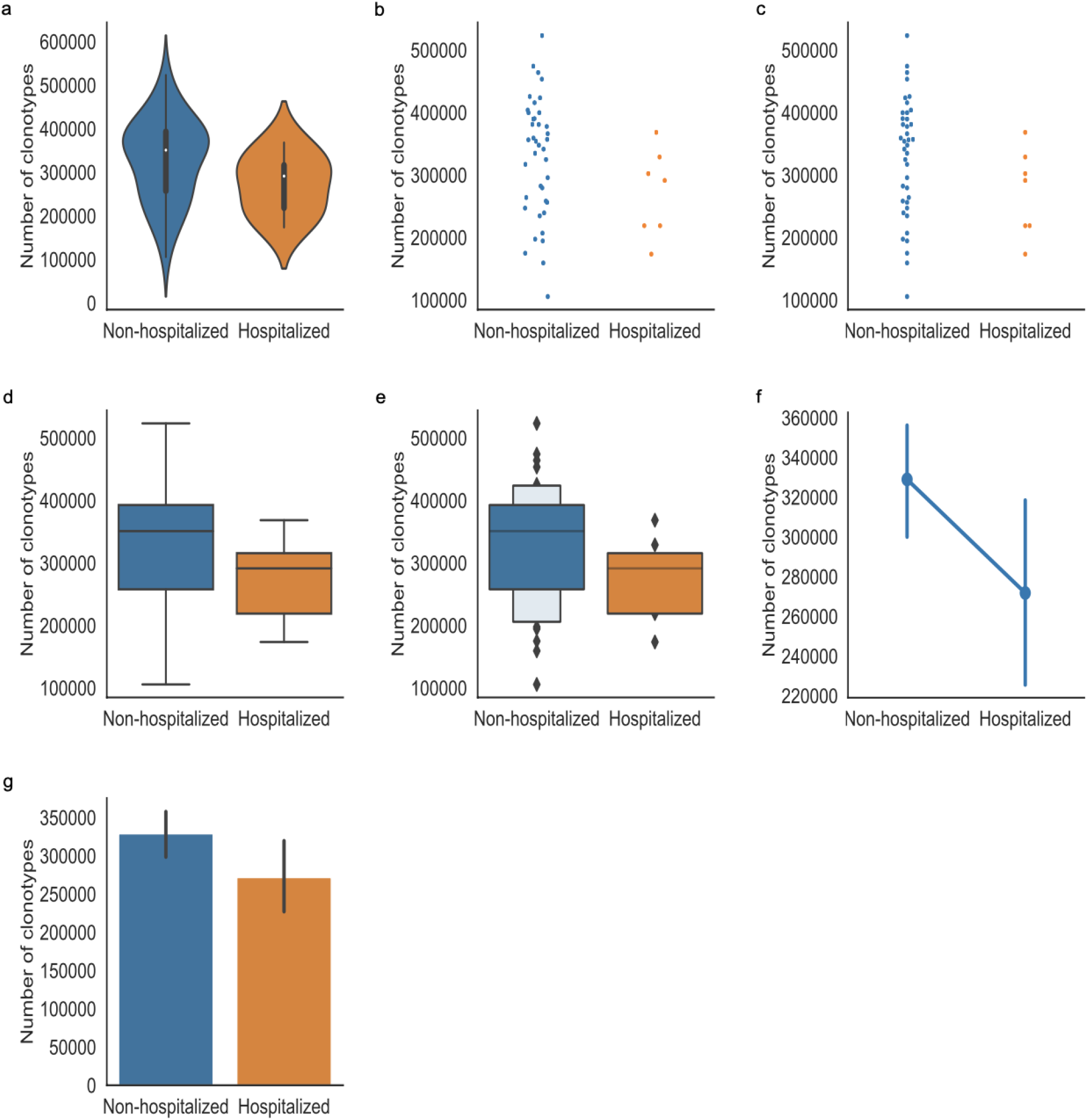
The clonotype counts were shown in groups by hospitalization status. The patients that were not hospitalized were shown in blue while the patients that were hospitalized were shown in orange. Different types of plots were shown as follows: (a) violin plot (b) strip plot (c) swarm plot (d) box plot (e) boxen plot (f) point plot (g) bar plot.

**Supplementary Fig.2.**
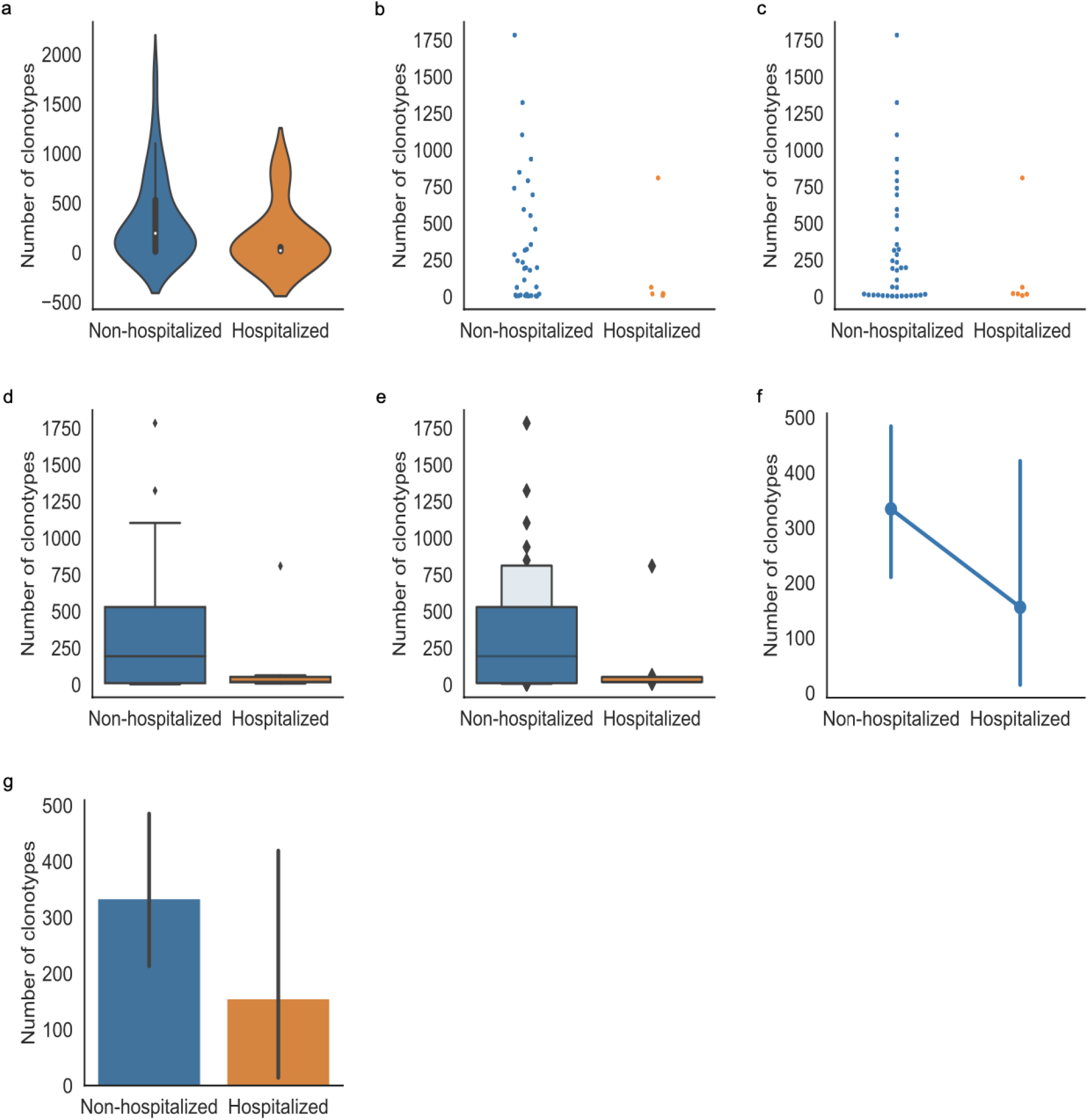
Clonal portion grouped by hospitalization status. The y-axis presented the number of clonotypes that counted for 10% of all the clonotypes in the repertoire. The patients that were not hospitalized were shown in blue while the patients that were hospitalized were shown in orange. Different types of plots were shown as follows: (a) violin plot (b) strip plot (c) swarm plot (d) box plot (e) boxen plot (f) point plot (g) bar plot.

**Supplementary Fig.3.**
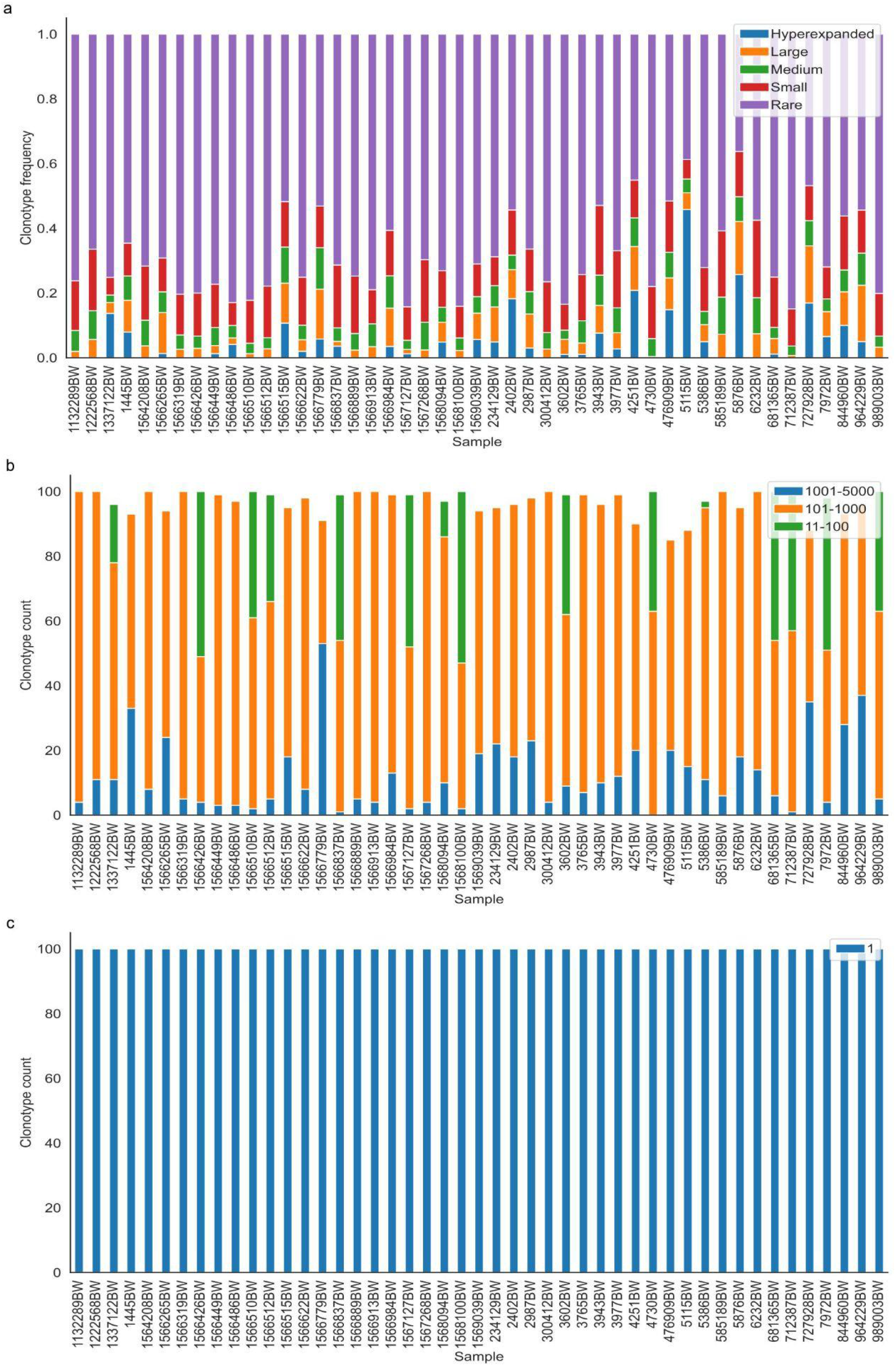
The distribution of clonotype groups in each sample. (a) The distribution of clonotype groups was based on the clonotype frequency across all the clonotypes. Hyperexpanded clonotypes (blue) were the clones with frequencies between 0.01 to 1, large clonotypes (orange) were the clones with frequencies between 0.001 to 0.01, medium clonotypes (green) were the clones with frequencies between 0.0001 to 0.001, small clonotypes (red) were the clones with frequencies between 0.00001 to 0.0001, rare clonotypes (purple) were the clones with frequencies between 0 to 0.00001. (b) The distribution of clonotype groups based on the clonotype count across the top 100 clonotypes. The clonotypes with clone counts between 1001-5000 were presented in blue, the clonotypes with clone counts between 101-1000 were resented in orange, and the clonotypes with clone counts between 11-100 were presented in green. (c) The distribution of clonotype groups based on the clonotype count across the rare 100 clonotypes. The clonotypes with clone count of 1 were presented in blue.

**Supplementary Fig.4.**
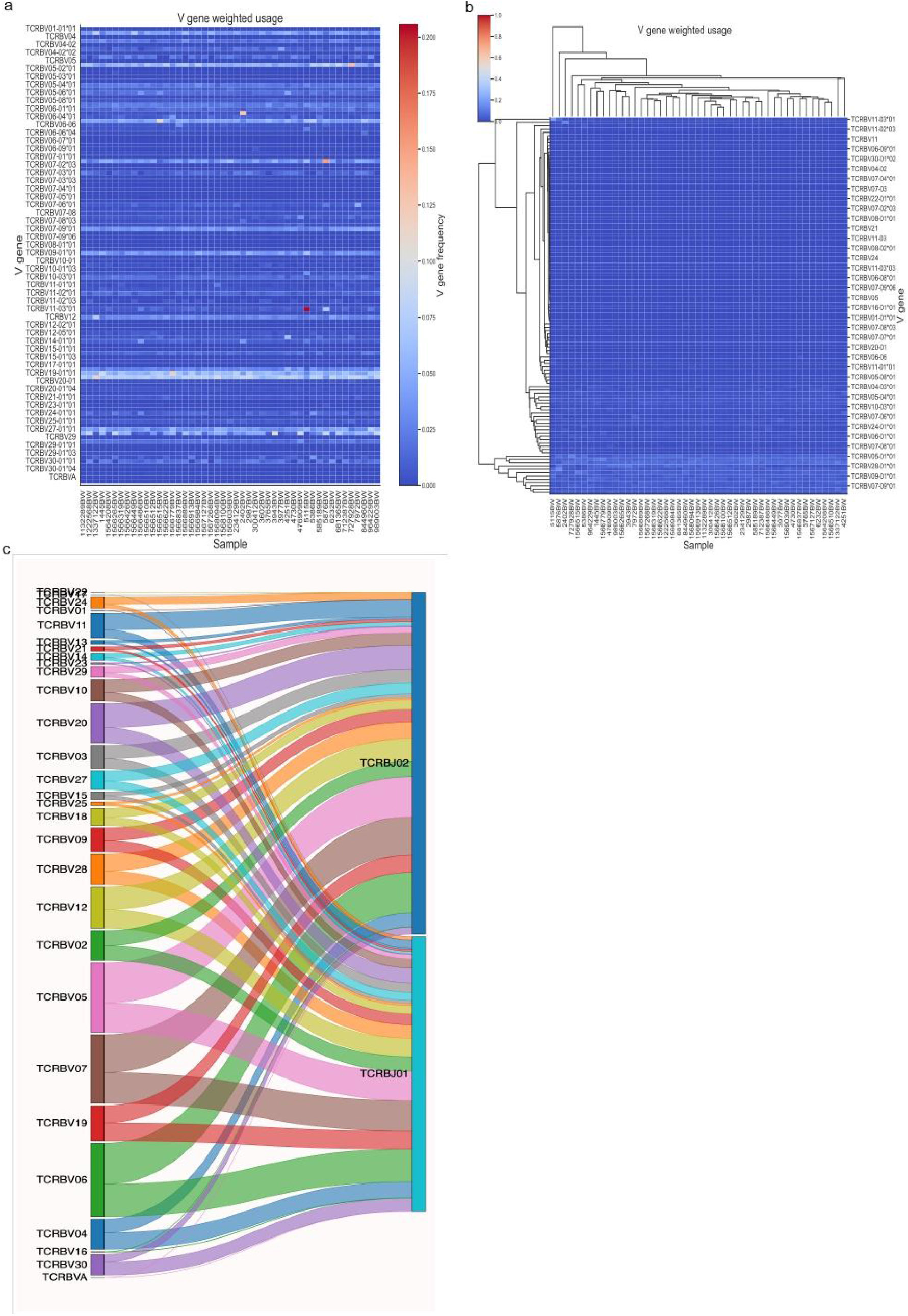
Heatmap of the weighted V gene usage in each sample and the Sankey plot for V-J combinations. (a) The heatmap of V gene weighted usage (b) The hierarchically-clustered heatmap of V gene weighted usage. x-axis represented each sample, y-axis represented different V genes. The shade of the color corresponded to the V gene frequency. (c) The V-J combinations with V and J gene frequencies in sample 3602BW were presented as a Sankey plot.

**Supplementary Fig.5.**
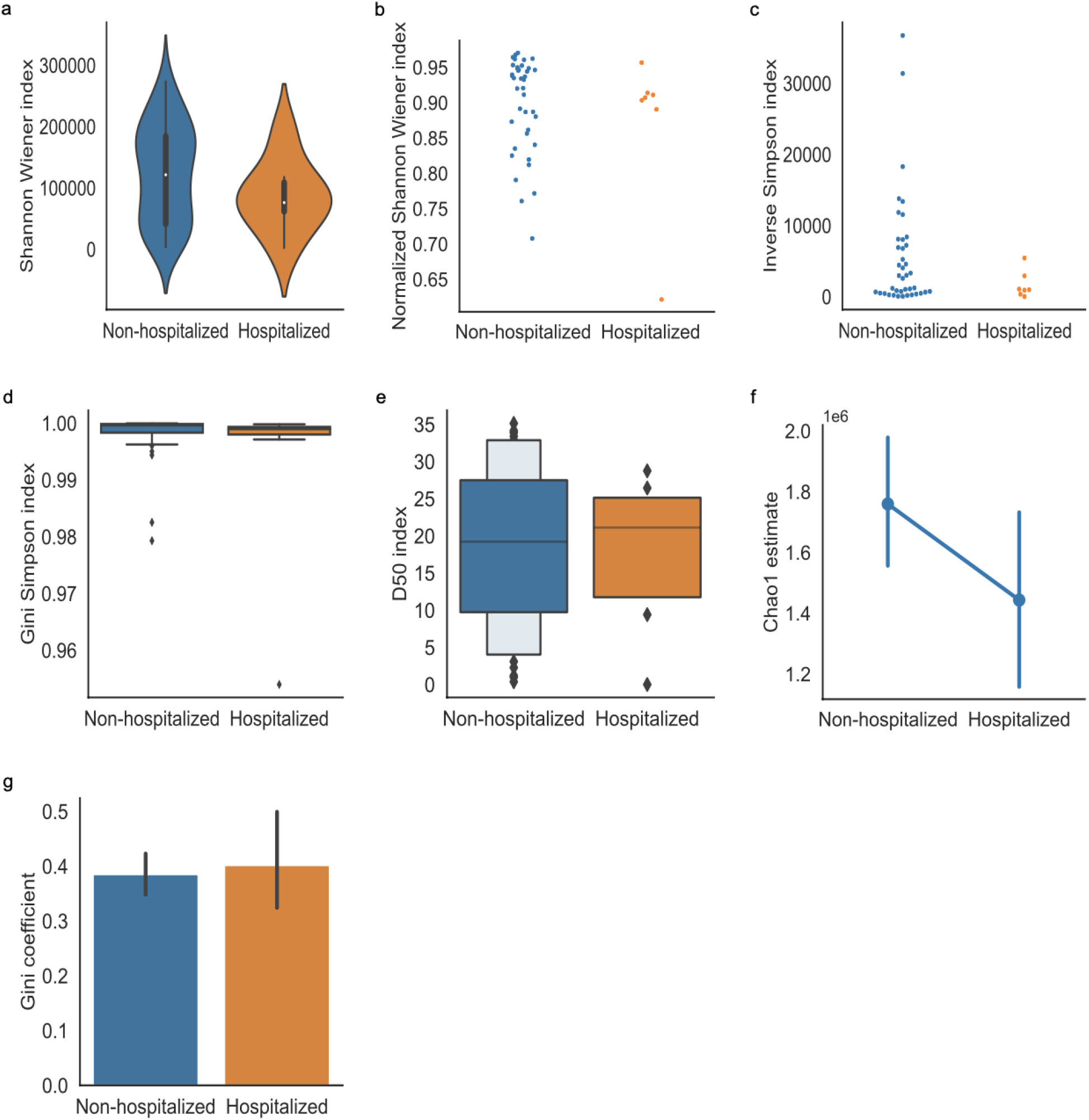
The diversity indices were shown in groups by hospitalization status. The patients that were not hospitalized were shown in blue while the patients that were hospitalized were shown in orange. (a) violin plot of the Shannon Wiener index (b) strip plot of normalized Shannon Wiener index (c) swarm plot of inverse Simpson index (d) box plot of Gini Simpson index (e) boxen plot of D50 index (f) point plot of Chao1 estimate (g) bar plot of Gini coefficient.

**Supplementary Fig.6.**
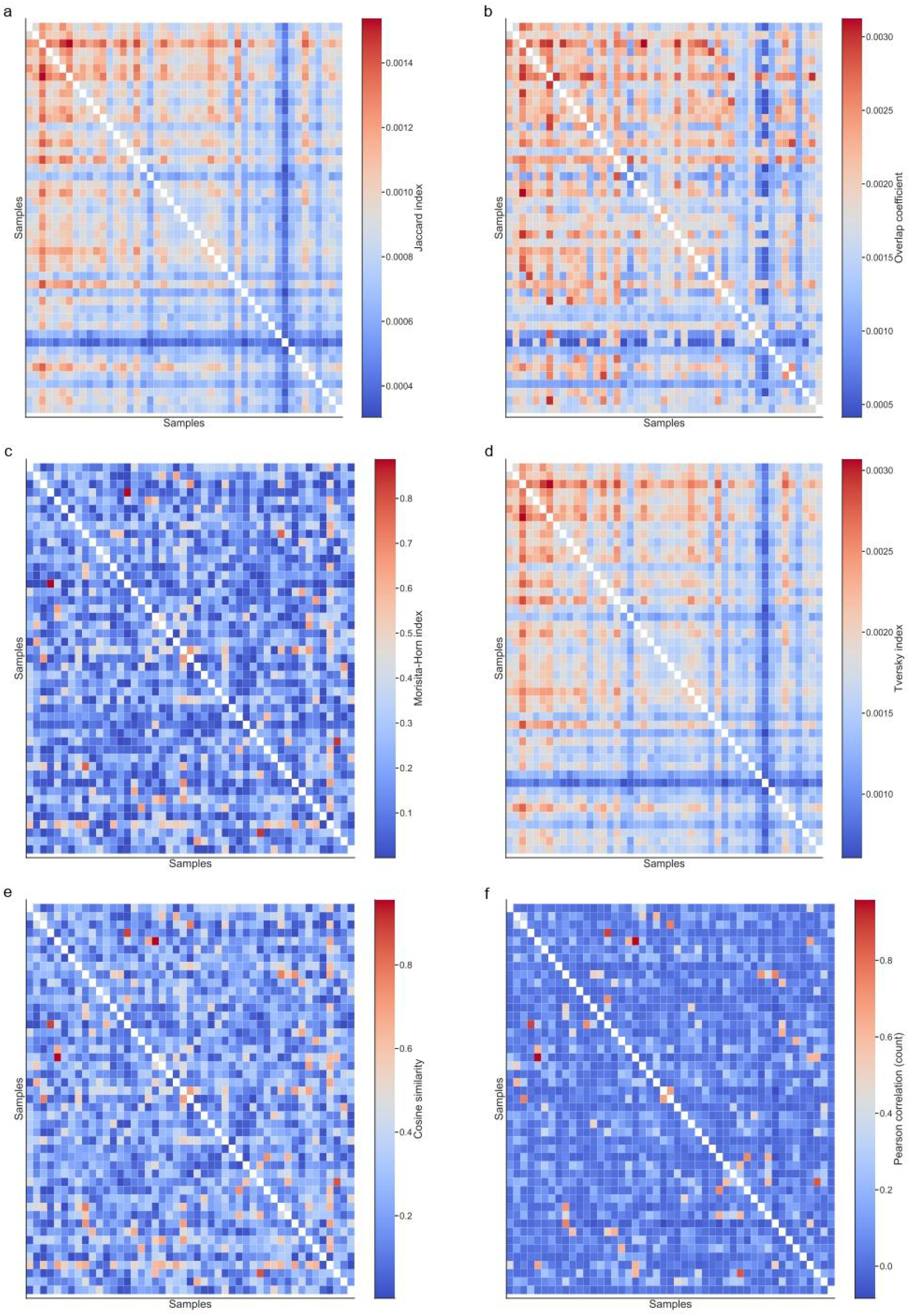
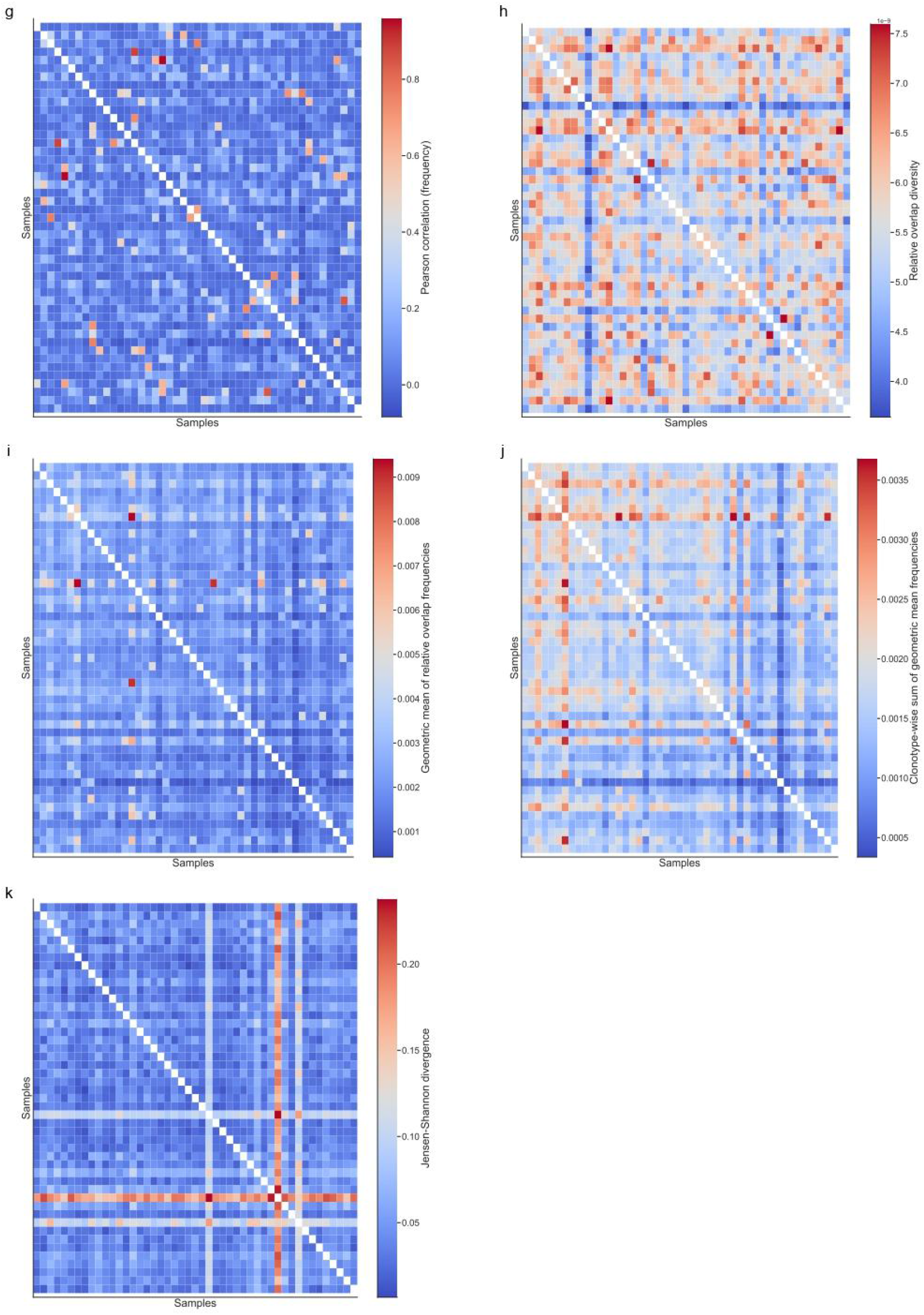
The overlap indices across samples. Heatmaps of each overlap index as shown above. The x-axis and the y-axis presented each sample. (a) Jaccard index (b) Overlap coefficient (c) Morisita-Horn index (d) Tversky index (e) Cosine similarity (f) Pearson correlation based on clonotype counts (g) Pearson correlation based on clonotype frequency (h) Relative overlap diversity (i) Geometric mean of relative overlap frequencies (j) Clonotype-wise sum of geometric mean frequencies (k) Jensen-Shannon divergence of variable gene usage distributions.

**Supplementary Fig.7.**
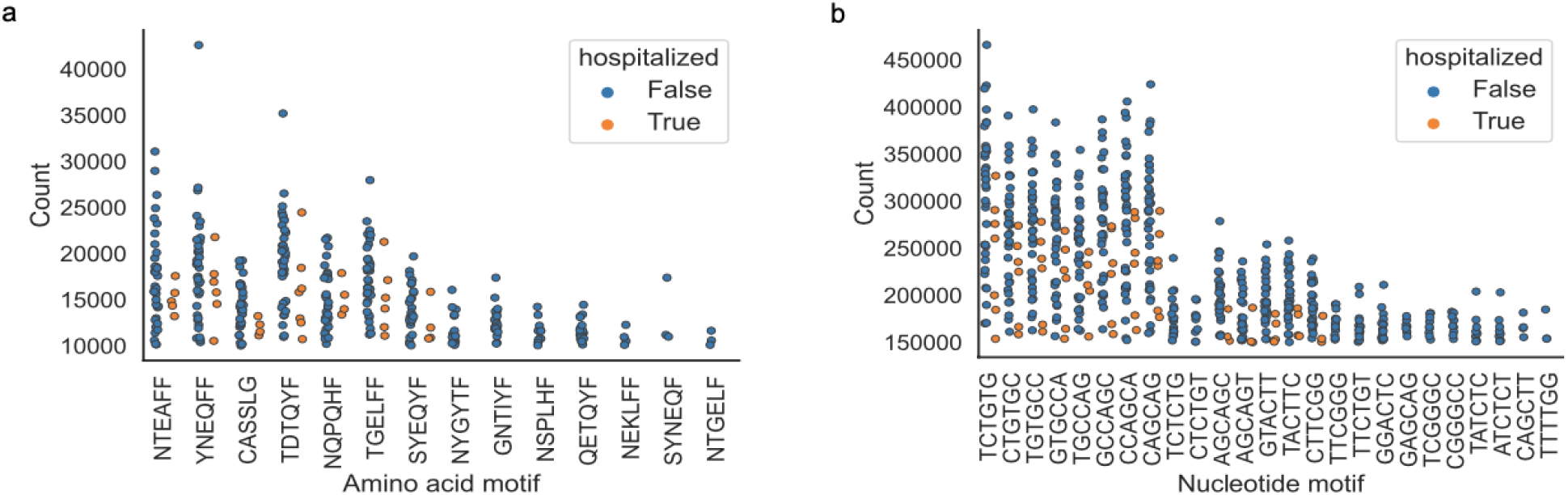
The number of highly presented motifs across samples grouped by hospitalization status. The x-axis presented motifs and the y-axis presented the count of the motifs. (a) amino acid motifs (k=6) that had numbers of counts of no less than 9,999 and were presented in more than 2 samples in hospitalization and non-hospitalization groups were shown in the strip plot (b) nucleotide motifs (k=6) that had numbers of counts of more than 150,000 and were presented in more than 2 samples in hospitalization and non-hospitalization group were shown in the strip plot.

## References

1. Aoki, H., Shichino, S., Matsushima, K. & Ueha, S. Revealing Clonal Responses of Tumor-Reactive T-Cells Through T Cell Receptor Repertoire Analysis. Front. Immunol. 13, 807696 (2022).

2. Liu, X. S. & Mardis, E. R. Applications of Immunogenomics to Cancer. Cell 168, 600–612 (2017).

3. Simnica, D. et al. High-Throughput Immunogenetics Reveals a Lack of Physiological T Cell Clusters in Patients With Autoimmune Cytopenias. Front. Immunol. 10, 1897 (2019).

4. Liu, X. et al. T cell receptor β repertoires as novel diagnostic markers for systemic lupus erythematosus and rheumatoid arthritis. Ann. Rheum. Dis. 78, 1070–1078 (2019).

5. Emerson, R. O. et al. Immunosequencing identifies signatures of cytomegalovirus exposure history and HLA-mediated effects on the T cell repertoire. Nat. Genet. 49, 659–665 (2017).

6. Luo, L. et al. Dynamics of TCR repertoire and T cell function in COVID-19 convalescent individuals. Cell Discov. 7, 89 (2021).

7. Gate, D. et al. Clonally expanded CD8 T cells patrol the cerebrospinal fluid in Alzheimer’s disease. Nature 577, 399–404 (2020).

8. Patas, K. et al. T Cell Phenotype and T Cell Receptor Repertoire in Patients with Major Depressive Disorder. Front. Immunol. 9, 291 (2018).

9. Hogan, S. A. et al. Peripheral Blood TCR Repertoire Profiling May Facilitate Patient Stratification for Immunotherapy against Melanoma. Cancer Immunol. Res. 7, 77–85 (2019).

10. Schrama, D., Ritter, C. & Becker, J. C. T cell receptor repertoire usage in cancer as a surrogate marker for immune responses. Semin. Immunopathol. 39, 255–268 (2017).

11. Shugay, M. et al. VDJtools: Unifying Post-analysis of T Cell Receptor Repertoires. PLoS Comput. Biol. 11, e1004503 (2015).

12. Nazarov, V., immunarch.bot & Rumynskiy, E. immunomind/immunarch: 0.6.5: Basic single-cell support. (Zenodo, 2020). doi:10.5281/zenodo.3893991.

13. Mangul, S. et al. Challenges and recommendations to improve the installability and archival stability of omics computational tools. PLoS Biol. 17, e3000333 (2019).

14. Randles, B. M., Pasquetto, I. V., Golshan, M. S. & Borgman, C. L. Using the Jupyter Notebook as a Tool for Open Science: An Empirical Study. in 2017 ACM/IEEE Joint Conference on Digital Libraries (JCDL) 1–2 (2017). doi:10.1109/JCDL.2017.7991618.

15. Chang, C.-M. et al. Profiling of T Cell Repertoire in SARS-CoV-2-Infected COVID-19 Patients Between Mild Disease and Pneumonia. J. Clin. Immunol. 41, 1131–1145 (2021).

16. Zhang, F. et al. Adaptive immune responses to SARS-CoV-2 infection in severe versus mild individuals. Signal Transduct. Target. Ther. 5, 156 (2020).

